# Factors Associated with Failure of Fecal Microbiota Transplant for Recurrent *Clostridioides difficile* Infection

**DOI:** 10.1101/2024.11.05.24316709

**Authors:** Joseph D. K. Nguyen, Kibret G. Yohannes, Initha Setiady, Emma C. Phillips, R. Ann Hays, Brian W. Behm, Cirle A. Warren, Jae Hyun Shin

## Abstract

**Background:** *Clostridioides difficile* infection (CDI) has emerged as a prevalent and recurrent antibiotic-associated infection. Fecal microbiota transplantation (FMT) is the most effective treatment for recurrent CDI (rCDI). Despite high success rates, FMT is ineffective in 5-20% of cases. Factors associated with failure have not been clearly defined. We seek to better understand factors predictive of FMT failure.

**Methods:** A retrospective chart review was conducted on adult patients who were screened at the Complicated *C. difficile* Clinic at the University of Virginia Health System and received FMT for rCDI between 2013 and 2022. Primary outcome was failure of FMT, defined as either rCDI or all-cause death within one year.

**Results:** 240 patients underwent FMT: 70.4% were female, median age was 68, and median episodes of CDI was 4. 24.6% experienced failure within the year (18.3% had rCDI and 7.1% died). Age 70 or older (p=0.007), male sex (p=0.013), ≥4 episodes of CDI (p=0.010), hypertension (p=0.010), diabetes mellitus (p=0.002), malignancy (p=0.034), high thyroid-stimulating hormone (p=2.696×10^−5^), anemia (p=0.002), and low zinc (p=0.025) were significantly associated with FMT failure on univariate analysis; age 70 or older (OR=2.66 [1.29-5.67]), ≥4 episodes of CDI (OR=3.13 [1.47-7.09]), and diabetes mellitus (OR=2.82 [1.25-6.50]) persisted to be associated with failure on multivariate analysis.

**Conclusions:** Our study shows that FMT remains an effective treatment for rCDI. We highlight several factors associated with FMT failure, such as older age, ≥4 episodes of CDI, anemia, elevated TSH, and low zinc, and the need for additional research to clearly define causality.

## Introduction

*Clostridioides (*formerly *Clostridium*) *difficile* infection (CDI) has emerged as a common antibiotic-associated infection in the United States. With nearly 500,000 American patients diagnosed with CDI annually, implications extend beyond individual health, significantly impacting infection control and healthcare expenditures [1–4]. 5-45% of patients with CDI experience recurrence, and in patients with recurrence, the risk of future recurrent CDIs (rCDI) increases to 60% [5–7]. The main mechanism of rCDI is thought to be through decreased microbiome diversity leading to increased susceptibility to relapse or reinfection with *C. difficile* [8]. Fecal microbiota transplantation (FMT), which aims to restore the physiologic diversity of intestinal microbiota and resist colonization, has emerged as the most effective therapy for rCDI [9]. With success rates of 80-95%, FMT has revolutionized rCDI treatment; guidelines recommend offering FMT to patients with rCDI who have failed antibiotic treatment [6, 10–13]. We have utilized FMT in our dedicated *C. difficile* clinic, which has outperformed antibiotic treatment, especially in patients who had 3 or more recurrent episodes [14].

Despite FMT’s high efficacy, FMT remains ineffective in 5-20% of cases [5, 10]. Prior studies have highlighted potential associations between antidepressant use [15, 16], cholecystectomy [17], inpatient status, pseudomembranes, immunocompromised status [18], and FMT failure, but definitive risk factors for FMT failure remain poorly understood [10]. In this study, we evaluate the efficacy of FMT in the treatment of rCDI and identify factors that may contribute to FMT failure.

## Methods

The research protocol was approved by the Institutional Review Board at the University of Virginia (IRB-HSR#23045, Fecal Microbiota Transplantation). As this is a descriptive study of patients who are undergoing treatment recommended by IDSA guidelines, no hazardous procedures or chemicals are involved. The privacy rights of human subjects have been observed.

### Study Setting, Study Design, and Data Collection

We conducted a retrospective cohort study of patients aged 18 or older evaluated for rCDI at the Complicated *C. difficile* Clinic (CCDC) at the University of Virginia Health System and treated with FMT between 2013 and 2022. The CCDC is a referral clinic attended by a multidisciplinary team including gastroenterologists and infectious diseases physicians. The interdisciplinary team held conferences, as needed, to review criteria and discuss patients who were immunocompromised or medically complex. Initial evaluation included an extensive history and comprehensive laboratory testing, detailed in Table 1, regardless of symptomatology or medical history. We had no clinical exclusion criteria; patients with underlying gastrointestinal (GI) disorders, such as inflammatory bowel disease (IBD) and irritable bowel syndrome (IBS), were not excluded if CDI episodes were appropriately documented. Electronic medical records were reviewed for demographic information, GI medical history, other medical history, surgical history, CDI history, labs within one month before FMT, and follow-up data. Data was entered into a secure REDCap (Research Electronic Data Capture) database.

**Table 1:** Laboratory studies ordered at initial visit.

| Laboratory studies | Stool studies |
| --- | --- |
| Complete blood count | *Campylobacter* |
| Comprehensive metabolic panel | *Cryptosporidium* |
| Thyroid stimulating hormone | *Giardia* |
| Vitamin A, B12, D and zinc | Norovirus |
| Immunoglobulin levels | Shiga toxin |
| Erythrocyte sedimentation rate and C-reactive protein | *Clostridioides difficile* toxin |
| Serologies for human immunodeficiency virus, cytomegalovirus, Epstein-Barr virus, human T-cell lymphotropic virus, syphilis, and *Strongyloides* | Culture |
| Tissue transglutaminase IgA | Ova and parasite |
|  | Lactoferrin |

### FMT Procedure and Patient Follow-Up

Patients were eligible for FMT if they had experienced three or more CDI episodes despite appropriate treatment. Donor stool was obtained from either a directed donor (family and friends, n=45; first cohort of patients) or universal donor (prescreened and purchased from OpenBiome, n=195 (Boston, MA, USA)). FMT was performed by depositing donor stool in colon via colonoscopy, and biopsies were obtained at the discretion of the gastroenterologist. FMT was repeated if needed until no pseudomembranes (indicative of CDI) were visualized on colonoscopy (Figure 1). After FMT, patients were instructed to contact the clinic if they had recurrent diarrhea or concern for rCDI.

**Figure 1:**
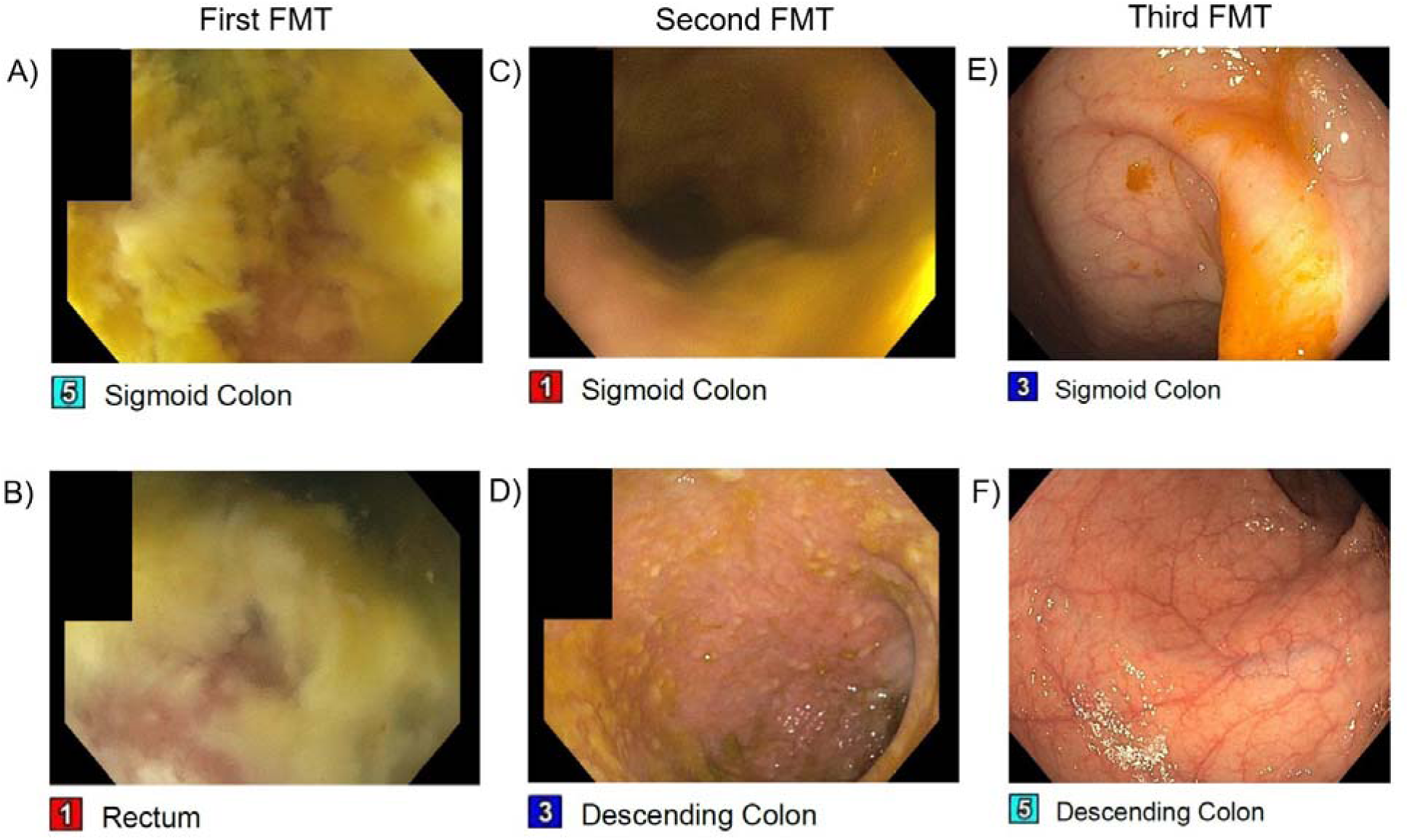
Colonoscopic findings from one patient shows edematous mucosa and pseudomembranes present throughout the sigmoid colon (A) and rectum (B) during the first FMT. On the second FMT (5 days after first FMT), there is some improvement in the sigmoid colon (C) and the scope was advanced to the descending colon which revealed still significant pseudomembranes (D). On the third FMT (2 weeks after first FMT), except for a congested proximal rectum, the rest of the examined colon, including both the sigmoid colon (E) and the descending colon (F) exhibited resolution of pseudomembranes and healthy intestinal epithelium.

### Study Outcomes

The primary outcome was FMT failure, defined as recurrence of CDI or death from any cause within one year of FMT (or both outcomes in series). Recurrence was defined as the return of diarrhea (liquid/loose stool ≥3 per day) with a stool sample testing positive for *C. difficile*. Though recurrence may occur multiple times, only the first instance was counted. Prior to February 2020, testing for *C. difficile* at our institution was initially done by toxin B gene polymerase chain reaction (PCR) (Xpert Cepheid, CA, USA); it was subsequently changed to a stepwise testing protocol consisting of multistep toxin B gene PCR with reflex, if PCR positive, to *C. difficile* toxin B enzyme immunoassay, with both results submitted to the treating clinician. Testing outside our institution was not uniform, but CDI episodes were recorded based on the clinical diagnostic decision of the treating physician. Recurrence and death were divided into early (<3 months after FMT) and late (3-12 months after FMT).

### Statistical Analysis

Chi-square analysis and t-test were used for univariate analysis, and multivariate binomial logistic regression model was used for multivariate analysis. Variables analyzed were age, sex, CDI, psychiatric conditions, use of biologics, prior cholecystectomy, hypertension, diabetes mellitus, GERD, hyperlipidemia, malignancy, immunosuppression, IBS, Crohn’s disease, ulcerative colitis, and diverticulitis. Laboratory values were not included in multivariate model due to incomplete documentation. All tests were two-tailed, and a p-value cutoff of 0.05 was used. Statistical analyses were performed with R, version 4.1.2.

## Results

240 patients with rCDI were treated with FMT at our facility from 2013 to 2022. Baseline characteristics of patients are reported in Table 2. 70.4% of patients were female and median age was 68 years (range: 20-96), with 47.5% of patients 70 or older. The median number of reported CDIs was four (range: 1-25), with 62.1% of patients experiencing four or more infections. 14.8% of patients had more than one FMT. 24.6% of patients experienced FMT failure, 18.3% of patients experienced recurrence, and 7.1% of patients died within one year of FMT (Figure 2A). Of the 18 patients who died, eight had a documented cause of death: failure to thrive (2), end-stage renal disease (2), infection (2), malignancy (1), and ileus (1).

**Figure 2:**
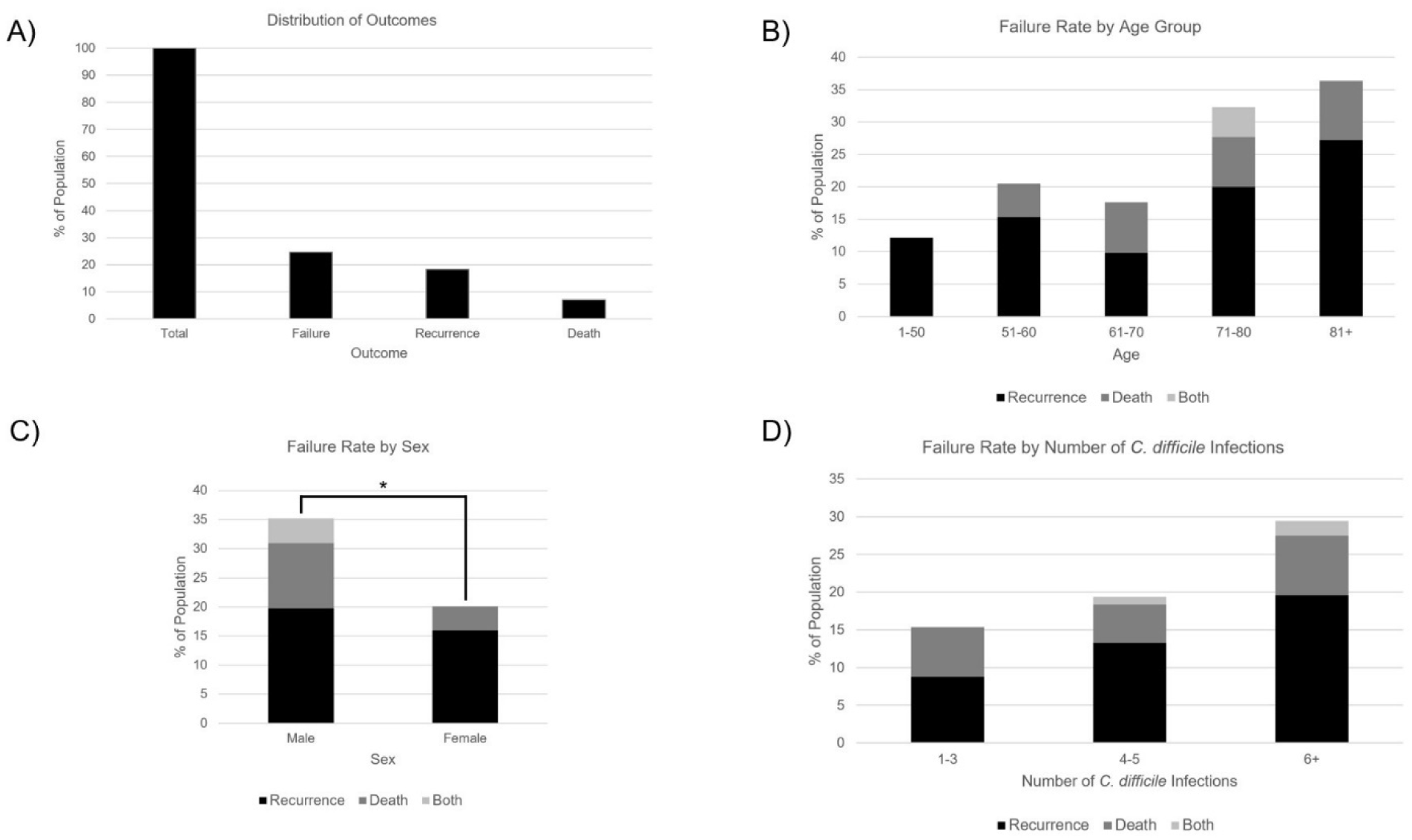
Rate of each adverse outcome (A), recurrence, death, and failure rate by age group (B), recurrence, death, and failure rate by sex compared with univariate analysis (C), and recurrence, death, and failure rate by number of *C. difficile* infections (D). No comparison was performed across groups in A, B, and D. Significance indicated by * (p<0.05), ** (p<0.01), or *** (p<0.001).

**Table 2:**
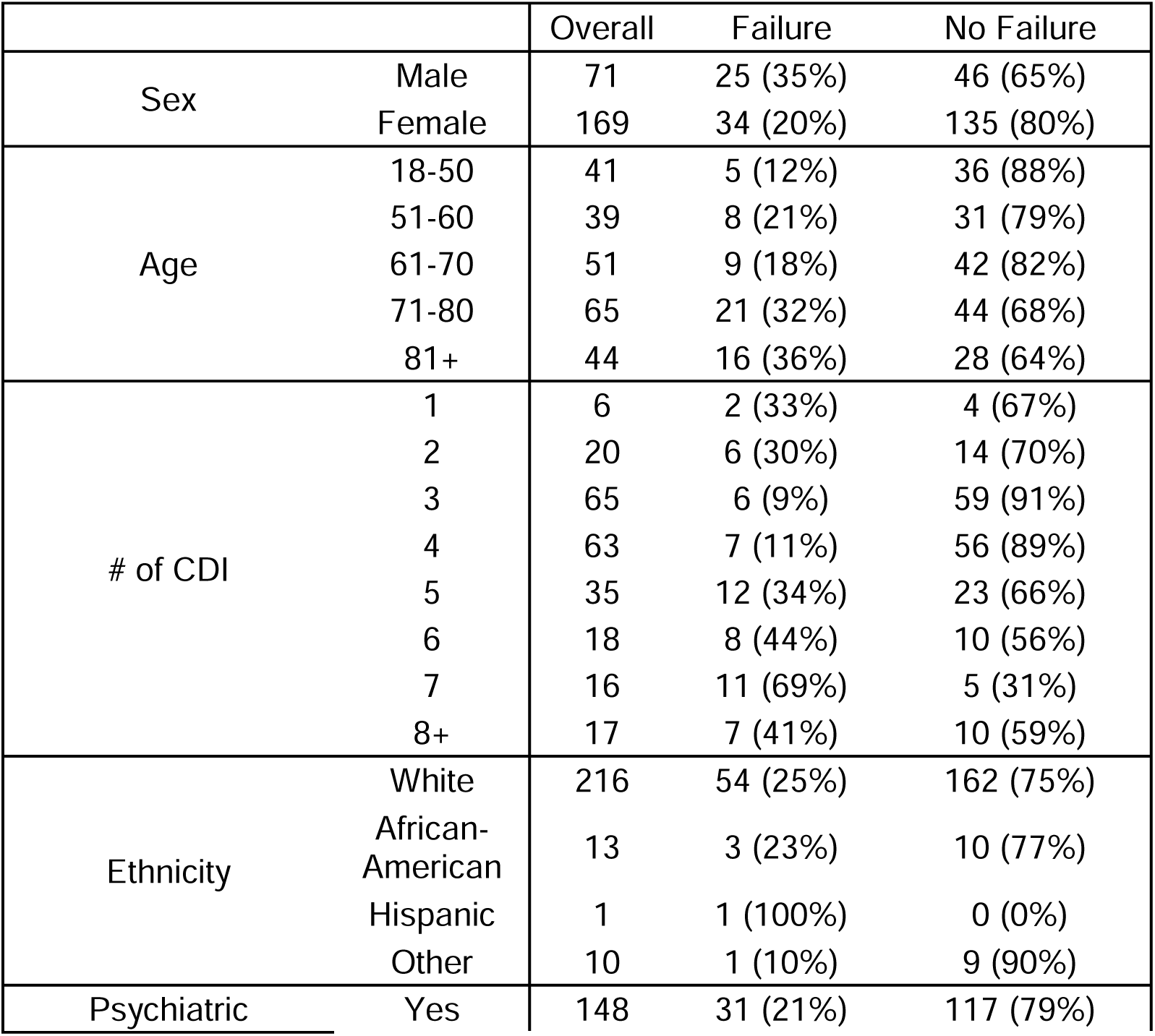
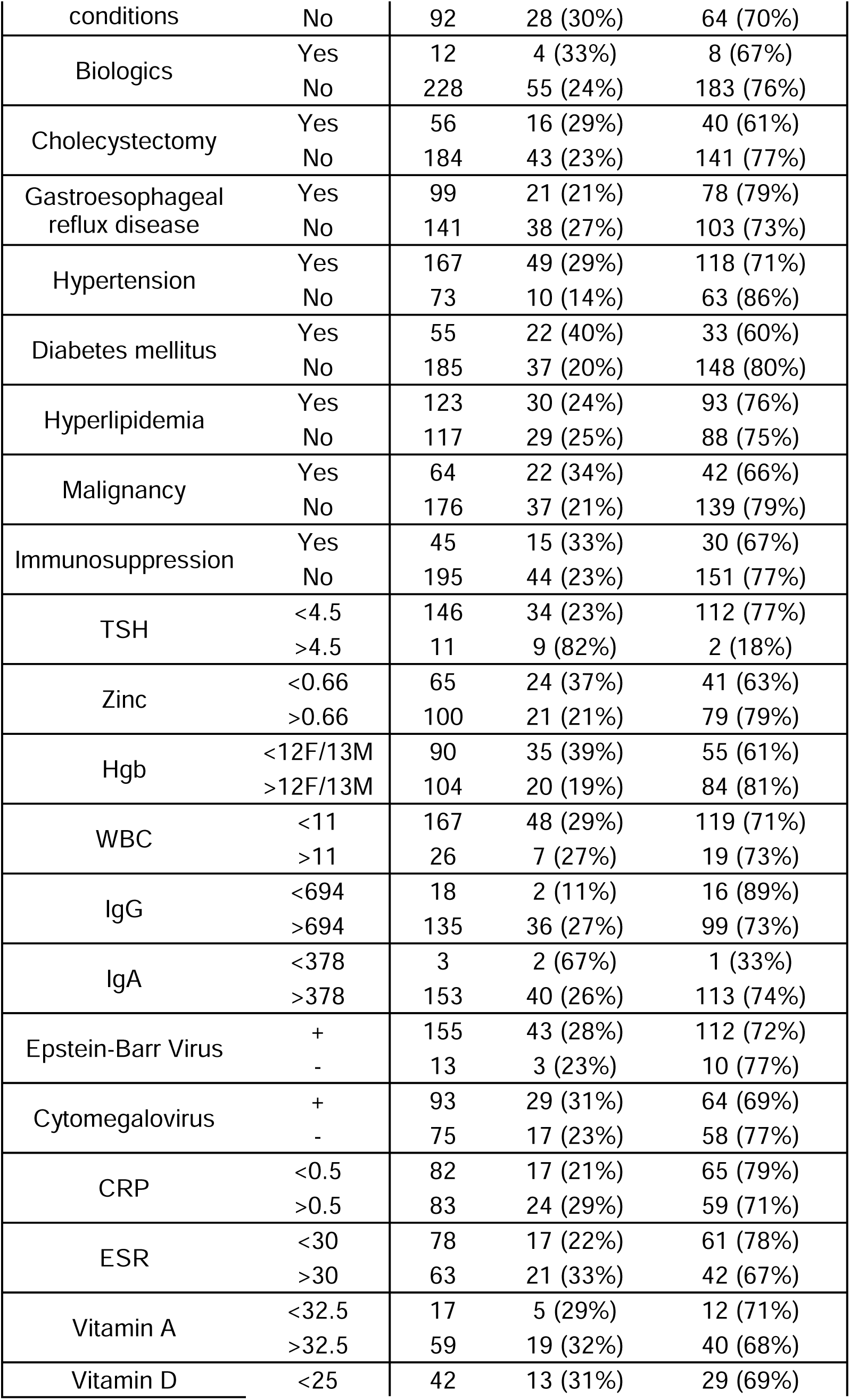
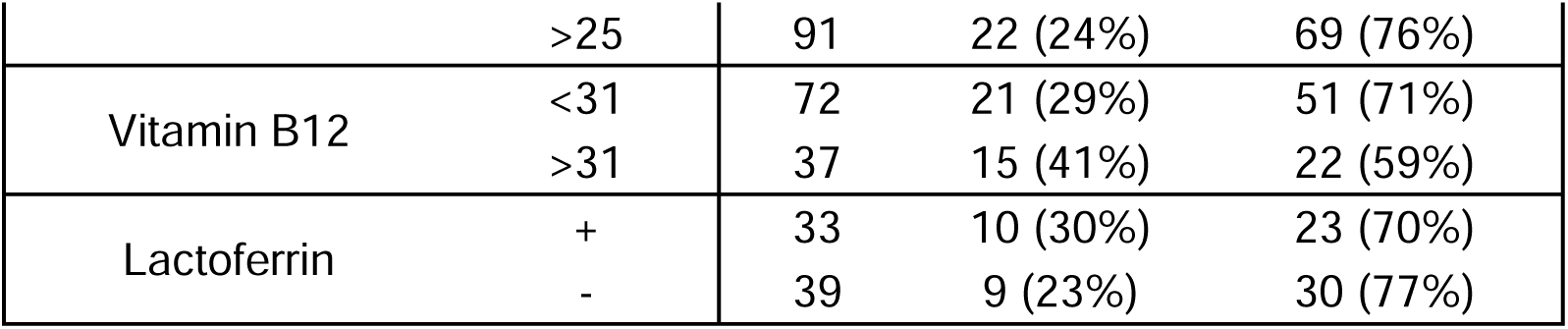
Demographics, comorbidities, and labs of patients by outcome.

In univariate analysis, age 70 or older (Figure 2B, 32% vs 17%, p=0.007), male sex (Figure 2C, 35% vs 14%, p=0.013), and four or more CDIs (Figure 2D, 30% vs 15%, p=0.010) were significantly associated with FMT failure. Regarding comorbidities, hypertension (29% vs 14%, p=0.010), diabetes mellitus (40% vs 20%, p=0.0025), and malignancy (34% vs 21%, p=0.034) were associated with FMT failure, while GERD (p=0.31), hyperlipidemia (p=0.94), and immunosuppressive conditions or medications (p=0.13) were not (Figure 3A). Ethnicity (p=0.24), psychiatric conditions (p=0.097), use of GABAergic psychiatric medications (p=0.97), use of non-GABAergic psychiatric medications (p=0.58), biologic treatments (e.g. monoclonal antibodies) (p=0.47), and prior cholecystectomy were also not associated with FMT failure (p=0.43). Regarding lab values, we found that high TSH (82% vs 23%, p=2.70e-5), low hemoglobin (Hgb, 39% vs 19%, p=0.0024), and low zinc (37% vs 21%, p=0.025) were associated with FMT failure (Figure 3B).

**Figure 3:**
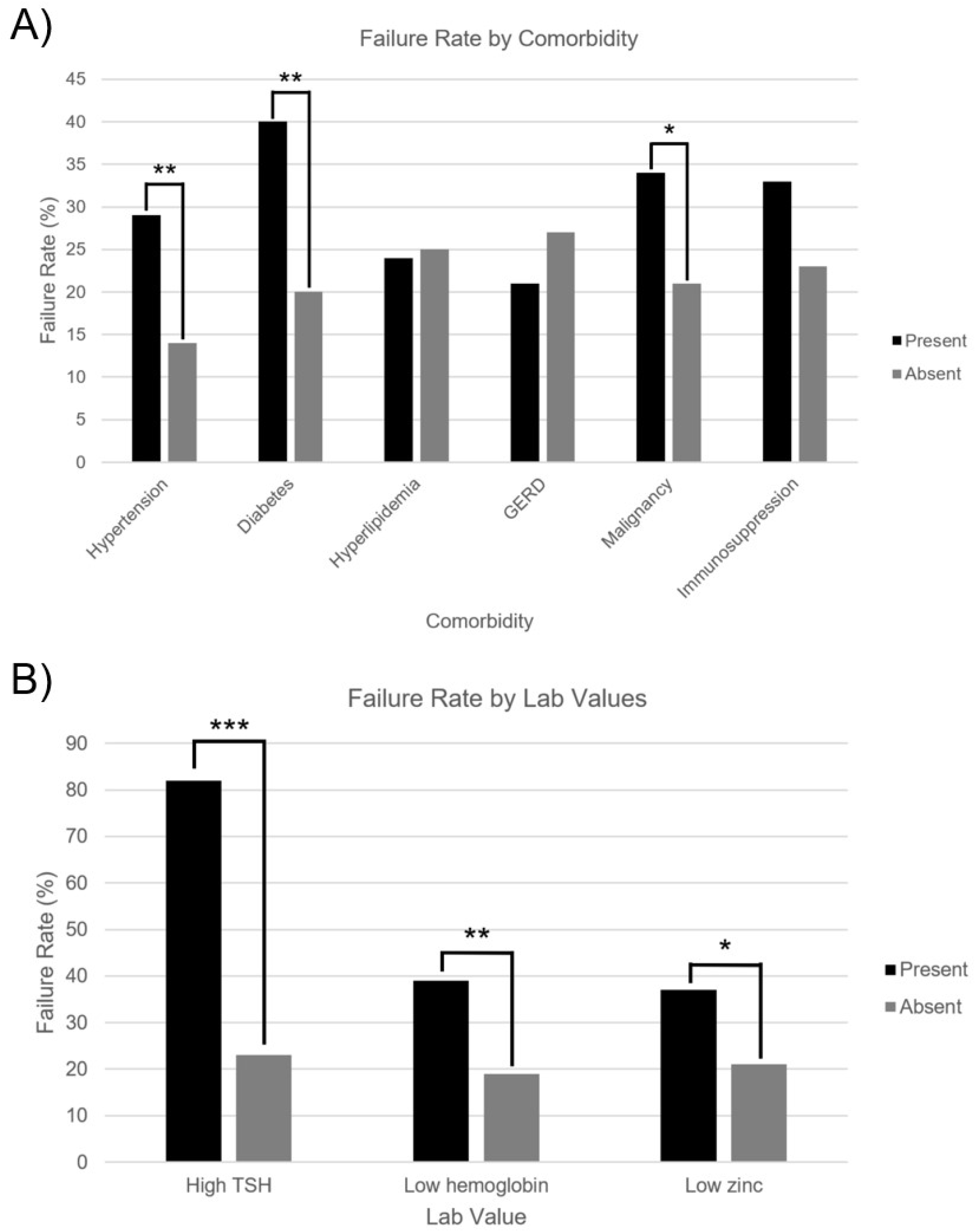
Univariate analysis of failure rate by presence vs absence of comorbidities (A) and abnormal vs normal lab values (B). Significance indicated by * (p<0.05), ** (p<0.01), or *** (p<0.001).

When breaking down failure into recurrence and death, recurrence was associated with age 70 or older (25% vs 13%, p=0.018), four or more CDI (24% vs 9%, p=0.0028), hypertension (22% vs 10%, p=0.021), and high TSH (63% vs 19%, p=6.35e-4). Death was associated with male sex (15% vs 4%, p=0.0023), diabetes mellitus (16% vs 5%, p=0.0045), high TSH (27% vs 5%, p=0.0032), low Hgb (17% vs 3%, p=0.00097), and low zinc (15% vs 3%, p=0.0039). No association was seen between recurrence or death and ethnicity, psychiatric conditions, cholecystectomy, GERD, or immunosuppression.

17% of patients experienced FMT failure early (within three months; 13.8% recurrence, 3.8% death). Of the 44 patients who recurred, 33 recurred early (75%) and 11 recurred late (25%). Age 70 or older (p=0.046), four or more CDI (p=0.03), hypertension (p=0.014), and high TSH (p=2.83e-4) were associated with early recurrence (compared to patients who recurred late or did not recur). Four or more CDI (p=0.029) continued to be associated with late recurrence (compared to patients who did not recur).

Multivariate analysis revealed age 70 or older (odds ratio (OR) 2.66 [1.29-5.67]), four or more CDI (OR 3.13 [1.47-7.09]), and diabetes mellitus (OR 2.82 [1.25-6.50]) were associated with failure while age 70 or older (OR 2.34 [1.08-5.24]) and four or more CDI (OR 3.63 [1.57-9.40]) were associated with recurrence.

## Discussion

rCDI remains a formidable challenge in clinical practice. Our cohort of 240 patients experienced an FMT success rate (i.e. no recurrence or death) of 83% over three months and 75% over one year, with a no-recurrence rate of 86% over three months and 82% over one year. This is in line with documented success rates (80-95%) when taking into account that most prior studies examined recurrence only (not death), and most only recorded recurrence within 1-3 months [10, 15, 19]. By expanding the study window and accounting for both recurrence and death, our study captures multiple factors that may not be seen with one outcome or a shorter study window. Nonetheless, our overall FMT success rate highlights the consistency and high efficacy of FMT in rCDI. Our study found the following factors to be correlated with either failure, recurrence or death: age 70 or older, male sex, four or more CDI, hypertension, diabetes mellitus, malignancy, high TSH, low hemoglobin, and low zinc.

Age over 70 is a well-documented risk factor for CDI and FMT failure, possibly due to comorbidities, including immunosuppression [20, 21]. However, even when controlling for comorbidities, age was an independent risk factor for FMT failure, a finding that has been corroborated in other studies [14]. Two hypotheses for this finding include: 1) age-related microbiome changes (shown to resemble antibiotic use in younger patients) [16, 19, 22], decreasing microbiome resistance to colonization and engraftment of the beneficial FMT microbiome and 2) deficient host immune response to FMT. Studies have shown that FMT can induce a global host immune response, possibly explaining its efficacy in treating rCDI [15]. This is mediated by innate lymphoid cell type 2 (ILC2) [16] and regulatory T cells (T_reg_) [19] through interferon-gamma [23] and Th17 responses [24]. Although an age-related deficiency in these mechanisms has not been demonstrated, analysis of immune responses in older patients in response to FMT would be helpful in elucidating these potential mechanisms.

Male sex was also associated with FMT failure in our cohort, particularly death. Previous studies have shown associations between female sex and FMT failure [25], female sex and initial CDI [11, 26], female sex and recurrent CDI [27], and male sex and death and severe complicated CDI [27]. Although only the last finding was seen in our cohort, it is notable that the proportion of women in the cohort was high (70.4%) and in line with previous studies (62%) [25]. Future directions include further analysis of potential confounders (e.g. increased antibiotic use in women due to more frequent urinary tract infections [30]) and bench studies regarding the mechanism of sex-related differences.

Medical comorbidities shown to be statistically significant factors in determining FMT failure included hypertension, diabetes mellitus, and malignancy, the first of which is a novel finding. On the other hand, diabetes mellitus has been shown to increase the risk of initial CDI [31] and rCDI [32]. While diabetes is associated with many infections, the association of both hypertension and diabetes mellitus may suggest an underlying metabolic contributor to FMT failure. Western diet, which is high in fat, has been associated with worse outcomes with CDI in animal models [20], while diet high in fiber was associated with a lower risk of CDI [33]. An exploration of diet in association with success or failure of FMT may be informative. Regarding malignancy, its primary contribution to failure was through increased deaths, although few malignancy-related deaths were explicitly documented. Malignancy has previously been associated with rCDI [34] and death from CDI [35–37]. Decreased immunity is thought to be an important contributor to this association [38], but an alteration of the microbiome by chemotherapy is also considered a possible mechanism [39]. Additionally, while psychiatric conditions [40] and immunosuppression (conditions or medications) [31, 32, 33] have previously been associated with FMT failure [40], we did not observe these relationships in our cohort.

Number of CDI episodes independently predicted FMT failure; specifically, patients with four or more episodes were more likely to have an unsuccessful FMT. While this may be reflective of a greater degree of dysbiosis in patients with more episodes of CDI [8], it also suggests that recurrence may be driven by factors not cured by FMT. Although it is unclear whether a higher number of CDI episodes directly contributes to FMT failure (as opposed to there being a different underlying cause for both increased CDI episodes and FMT failure), this finding suggests that lowering the threshold for switching from antibiotic treatment to FMT may be beneficial in some patients [41].

In addition to comorbidities, we studied lab values within 1 month prior to FMT. Low Hgb has been linked to FMT failure, a trend reaffirmed in this study [9]. Elevated ESR, an inflammatory marker, was numerically higher in patients in our cohort with FMT failure. The combination of low Hgb and high ESR may suggest the presence of anemia of chronic disease, a potential indicator of chronic inflammation. Chronic inflammation may be either a symptom of dysbiosis [42] or the cause of dysbiosis [43], or both, which may contribute to FMT failure.

Previous studies have indicated a correlation between zinc deficiency and FMT failure, which was seen in our cohort [29]. Patients afflicted with rCDI are susceptible to zinc deficiency due to diarrhea and anorexia associated with active CDI. Zinc has been shown to have antidiarrheal effects due to its ability to enhance water and electrolyte absorption, which aids in mucosal integrity, brush border enzyme activity, and immunity. Zinc has also been shown to play a potentially important role in sustaining a diverse microbiome [29]. Finally, decreased zinc levels may be a marker of intestinal inflammation, which may contribute to FMT failure.

A novel finding that emerged from this study is the association between elevated TSH and FMT failure. It is unclear whether elevated TSH reflects true clinical hypothyroidism, as reflex free T4 levels were rarely obtained in our cohort. In the existing literature, case reports have shown an association between CDI and decreased TSH, but not increased TSH [27, 28]. We present several hypotheses for this association. First, hypothyroidism is known to decrease gut motility. While this may be beneficial after FMT to increase the effects of treatment and maximize interaction between the donor stool and the recipient microbiota [26], it may worsen CDI and contribute to recurrence by retaining bacteria and toxins. Second, it is possible that elevated TSH reflects a hypothyroid phase in thyroiditis, although this is less likely due to its transient nature (while patients receiving FMT are chronically ill). Third, elevated TSH may be a manifestation of other underlying conditions associated with FMT failure. Finally, advanced age may contribute to elevated TSH; in our cohort, patients with normal or low TSH had a median age of 68, while patients with high TSH had a median age of 75.

Strengths of this study include: 1) consistent and standardized procedures and documentation, as all patients received FMT and an infectious diseases consultation at our institution, and 2) easily quantifiable endpoints, such as recurrence/death. Limitations of this study include: 1) possible confounding bias, stemming from unrecognized factors not controlled for, 2) clinically subjective indications for FMT and no documentation of CDI severity, leading to within-cohort differences that may be difficult to quantify, 3) incomplete documentation (i.e. not all patients received all pre-FMT blood tests), 4) lack of long-term follow-up beyond one year, and 5) non-uniform post-FMT management (e.g. no documentation on suppressive antibiotics).

In conclusion, our study has identified several factors associated with failure of FMT in the treatment of rCDI, including age over 70, male sex, four or more CDIs, hypertension, diabetes mellitus, malignancy, elevated TSH, low Hgb, and low zinc. FMT remains a highly efficacious treatment for rCDI; at one-year follow-up, about 3 in 4 patients who underwent FMT did not have recurrent CDI or death. Awareness of demographic and comorbid factors associated with FMT success can better inform care for patients undergoing FMT for rCDI. Future directions include expanding the cohort and exploring mechanisms underlying the associations uncovered in this study.

## Data Availability

All data produced in the present study are available upon reasonable request to the authors.

## Writing Assistance

The authors received no writing assistance.

## Author Contributions

E.P., C.A.W., and J.H.S. designed the study, J.N., I.S., K.Y., and E.C.P. performed data collection, J.N. performed statistical analysis, J.N., I.S., K.Y., C.A.W., and J.H.S. interpreted the data, J.N., I.S., and K.Y. drafted the manuscript, J.N., I.S., K.Y., E.C.P., R.A.H., B.W.B., C.A.W., and J.H.S. critically revised the manuscript for important intellectual content, J.H.S. and C.A.W. provided administrative support and supervised the study.

## Glossary

CBC: complete blood count
CCDC: Complicated *Clostridioides difficile* Clinic
CDI: *Clostridioides difficile* infection
CMP: complete metabolic panel
CRP: C-reactive protein
ESR: erythrocyte sedimentation rate
FMT: fecal microbiota transplantation
GERD: gastroesophageal reflux disease
GI: gastrointestinal
IBD: inflammatory bowel disease
IBS: irritable bowel syndrome
ILC2: innate lymphoid cell type 2
OR: odds ratio
PCR: polymerase chain reaction
rCDI: recurrent *Clostridioides difficile* infection
REDCap: Research Electronic Data Capture
T_reg_: regulatory T cell
TSH: thyroid-stimulating hormone
WBC: white blood cell count.

